# A randomized crossover trial of short versus conventional pulse width DBS in Parkinson’s Disease

**DOI:** 10.1101/2021.06.20.21258955

**Authors:** Jan Niklas Petry-Schmelzer, Lisa M Schwarz, Hannah Jergas, Paul Reker, Julia K. Steffen, Haidar S. Dafsari, Juan Carlos Baldermann, Gereon R. Fink, Veerle Visser-Vandewalle, Till A. Dembek, Michael T. Barbe

## Abstract

**Background:** Subthalamic nucleus deep brain stimulation is a well-established treatment for patients with Parkinson’s disease. Previous acute challenge studies suggested that short pulse widths might increase the therapeutic window while maintaining motor symptom control.

**Objectives:** To investigate in patients with Parkinson’s disease and nucleus subthalamicus deep brain stimulation (STN-DBS) whether short pulse width stimulation with 30µs maintains equal motor control as conventional 60µs stimulation over a period of 4 weeks.

**Methods:** In this monocentric, double-blinded, randomized crossover trial, 30 patients with Parkinson’s disease and STN-DBS were enrolled and assigned to 4 weeks of stimulation with 30µs and 4 weeks of stimulation with 60µs in randomized order (German Clinical Trials Register No. DRKS00017528). The primary outcome was the difference in motor symptom control as assessed by a motor diary. Secondary outcomes included energy consumption measures, non-motor effects, side-effects, and quality of life.

**Results:** A total of 24 patients were included in the final analysis. There was no difference in motor symptom control between the two treatment conditions. Concerning secondary outcomes there was no difference in energy consumption, non-motor symptoms, side-effects, or quality of life. On the individual level, patients preferring 30µs tended to be more dyskinetic in the 60µs setting, whereas patients preferring 60µs experienced more off-time in the 30µs setting.

**Conclusions:** Short pulse width settings (30µs) provide equal motor symptom control as conventional (60µs) stimulation without significant differences in energy consumption. Future studies are warranted to evaluate a potential benefit of short pulse width settings in patients with pronounced dyskinesia.

## Introduction

Subthalamic nucleus deep brain stimulation (STN-DBS) is a well-established treatment for patients with advanced Parkinson’s disease (PD) improving quality of life, motor, and non-motor symptoms.^1–3^ However, postoperative outcomes critically depend on lead locations and stimulation settings.^4–7^ While titration of the stimulation amplitude and choice of the active contact are usually individual, stimulation frequency and pulse width are typically set to standard values of 130Hz and 60µs. This standard value emerged from a previous study, demonstrating that a pulse width of 60µs enlarges the therapeutic window.^8,9^

New generations of DBS systems enable the use of even shorter pulse widths per clinical routine. Several recent studies suggested that shorter pulse widths, i.e. below 60µs, might lead to an additional widening of the therapeutic window, defined as the amplitude range between threshold amplitudes for rigidity control and occurrence of side-effects.^10–14^ Additionally, some authors suggested that DBS with shorter pulse widths might reduce energy consumption while maintaining equal symptom control compared to standard pulse width as investigated during an acute challenge design.^10–12^ Furthermore, DBS with 30µs resulted in an improvement of stimulation-induced dysarthria, dyskinesia, and gait in an acute challenge^14,15^. This beneficial effect on stimulation-induced dysarthria and gait was neither replicated in a double-blinded randomized crossover trial investigating a period of 4 weeks in a cohort of PD patients with stimulation-induced dysarthria,^13^ nor in a recent study investigating the effect of short pulse width stimulation on objective gait parameters in an acute challenge.^16^ As a secondary outcome of these study by Dayal et al., there was no difference in motor symptom control between short and conventional pulse width stimulation, captured as a snapshot by the Movement Disorder Society-Unified Parkinson’s Disease Rating Scale.^13^

To summarize, shorter pulse widths were associated with the potential to reduce side-effects and energy consumption while maintaining the therapeutic effect. Whether these effects can be reconfirmed beyond the setting of an acute challenge, remains to be elucidated. Therefore, in this double-blind, randomized, crossover trial, we investigate whether short pulse width stimulation with 30µs maintains motor symptom control, as measured by a patient-reported motor diary, in comparison to 60µs for 4 weeks.

## Methods

### Participants and Ethical Approval

Eligible patients were aged 18 to 80 years with a clinical diagnosis of PD following the UK Brain Bank criteria, who underwent bilateral STN-DBS at least 3 months before study inclusion. Patients had been selected for STN-DBS according to the guidelines of the International Parkinson and Movement Disorders Society.^17^ Additionally, the implanted pulse generator required the possibility of providing stimulation with a pulse width of 30µs as per clinical routine. The local ethics committee approved the trial (vote: 19-1233) conducted under the declaration of Helsinki, which was registered with the German Clinical Trials Register (registration No. DRKS00017528). Data was collected at the Department of Neurology of the University Hospital Cologne.

### Study Design

This study was a single-center, randomized, double-blind, crossover trial of neurostimulation with a short pulse width (30µs) versus standard pulse width (60µs) in PD patients with bilateral STN-DBS. The trial protocol consisted of 3 visits per participant. At baseline, medical and surgical history, as well as the current medication were recorded. Stimulation settings with a pulse width of 60µs (PW60) and 30µs respectively (PW30) were established based on the optimized clinical setting at study inclusion in the “medication *ON”* state. Of note, the active electrode and frequency were equal for both trial programs, whereas the stimulation amplitude was titrated for the cardinal symptoms and side effects. Patients were trained to titrate the amplitude with the handheld patient programmer to perform adjustments of the stimulation amplitude within individual ranges if needed. The clinician tested the range of the stimulation amplitude to exclude acute stimulation-induced side effects. Then patients were randomized (blockwise (3 Blocks) 1:1 randomization using www.randomizer.at) to begin with PW60 followed by four weeks with PW30 or vice versa. No changes in medication were allowed throughout the trial.

After 4 weeks (Period A), patients returned for a clinical assessment after at least overnight withdrawal of dopaminergic medications (follow up visit1: 28 ±2 days from baseline). Adverse events and stimulation parameters, including impedances, were recorded. Secondary outcomes were assessed, and stimulation was then switched to the other treatment condition. If necessary, the stimulation amplitude was again optimized before discharge. Patients returned after 4 weeks (Period B) to repeat the assessment in “medication *OFF”* state (follow up visit 2: 56 ±2 days from baseline). By the end of the trial, patients were asked for their preferred setting. Besides the follow-up visits, a telephone call was done after 14 days of each period and 5 days prior to each visit to support the patient in self-adjustment of the amplitude if needed. If one treatment condition was not well tolerated despite adjusting the amplitude, patients could choose between an early cross-over or end of study after filling the motor diary over three days, or an instant termination of the trial. All participants and assessing clinicians remained blinded to the treatment condition throughout the trial. Unblinded clinicians programmed the stimulation settings.

### Outcome Parameters

The primary outcome parameter was the difference in a standardized self-reported motor diary between the PW60 and the PW30 condition over the last three days in each condition.^18^ The motor diary assesses the motor state defined as (1) awake and with good symptom control, (2) awake and troublesome dyskinesia, (3) awake and with poor motor function, and (4) asleep in intervals of 30 minutes. Previous studies have already used this diary to evaluate motor symptom control in PD patients with STN-DBS, beyond an acute assessment.^2,19,20^ For further analysis the mean time of the respective motor state in each period was calculated.

Predefined secondary outcomes, as assessed in “medication *OFF*” state at each follow-up visit, were differences in the Unified Parkinson’s Disease Rating Scale (UPDRS-I, -II and -III), 10-m timed walk test, visual analog scale (VAS) for self-assessment of “ability to walk”, and “ability to speak”, Parkinson’s disease questionnaire 39-Item Quality of Life Questionnaire Summary Index (PDQ-39 SI), and Non-Motor Symptoms Questionnaire (NMSQ). Additionally, differences in speech intelligibility were investigated by reading a German standard text (“Der Teppichklopfer”) rated by 13 blinded naïve listeners on a VAS. All VAS ranged from 0 to 10 (“worst possible state” to “best possible state” of the respective symptom). To investigate differences in energy consumption the battery charge index (BCI),^21^ the total charge delivered per pulse (CPP = current*pulse width), and the total electrical energy delivered (TEED = ((current*impedance)^2^*frequency*pulse width)/ impedance) were calculated, and the sum of both hemispheres was reported.

### Statistical Analysis

An intention-to-treat analysis was conducted for all outcome parameters employing linear mixed effects models including the grouping variable (PW60, PW30) as a fixed effect and subject as a random effect to correct for the paired nature of the data. For analysis of the primary outcome parameter the interaction between period and the grouping variable (PW60, PW30) was included as an additional fixed effect to account for a possible period effect. A carryover effect was not expected as there was a wash-out period between the two follow-up assessments. The power analysis for non-inferiority regarding the primary outcome parameter with a power of 80% and a significance level of 0.05 based on a previous trial by Timmermann et al.^19^ resulted in a total of 27 patients needed. When considering a drop-out rate of 10%, a total of 30 patients had to be recruited. For subgroup analysis (comparison of non-preferred settings) Shapiro-Wilk-tests were conducted to test for normal distribution. Then paired t-test or Wilcoxon signed rank test was employed, respectively. The Bonferroni method was used to correct for multiple comparisons and statistical significance was set to p < 0.05. Results are reported as mean and standard deviation if not indicated otherwise. We used MATLAB R2020a (The MathWorks Inc., Natick, Massachusetts, United States) for all data analysis.

## Results

### Participants

Between August 15th, 2019, and December 3rd, 2020, 30 patients were enrolled. A total of 24 patients were included in the intention-to-treat analysis (7 female) as 6 patients had to be excluded or dropped out of the trial during the first period of the trial. Figure 1 depicts the participant flow and reasons for dropout. Patients included in the analysis were 61.4 years (±8.7) old, had a mean disease duration of 8.8 years (±3.8), and were included 0.9 year (±0.8) after surgery. The median Hoehn&Yahr stage was 2.5 (IQR: ±1.0) and the mean levodopa equivalent daily dose at study inclusion was 443.5 mg (±214.2). Figure 2 shows the order of the pulse width settings throughout the trial and the preferred settings at the end of the trial. All patients were implanted with the same implantable pulse generator (Gevia™, Boston Scientific), and Cartesia™ leads (Boston Scientific) except one patient (Medtronic 3387 leads). Adverse events and stimulation settings are reported in the supplement (Supplementary Table 1 and Supplementary Table 2).

**Fig. 1.**
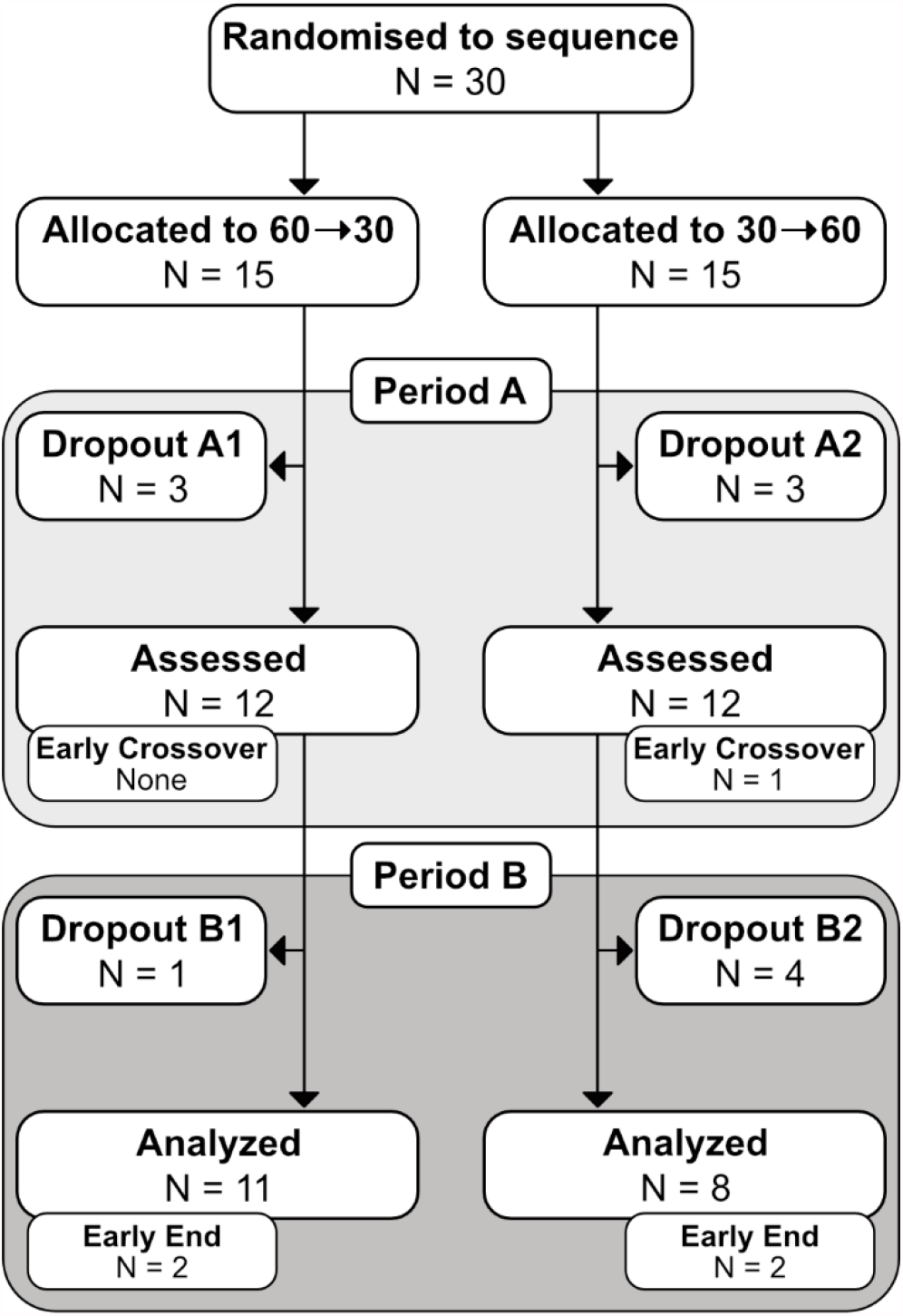
Participant Flow. Reasons for dropout were: Withdrawal of consent due to COVID-19 pandemic (N = 3; 1 A1, 1 A2, 1 B2), insufficient symptom control (N = 2; 1 A2, 1 B1), withdrawal of consent without specific reason (N = 3; 2 A1, 1 B2), DBS system infection (N = 1; B2), exclusion due to violation of study protocol (unauthorized switching of stimulation programs during the trial, N = 2; 1 A2, 1 B2)).

**Fig. 2.**
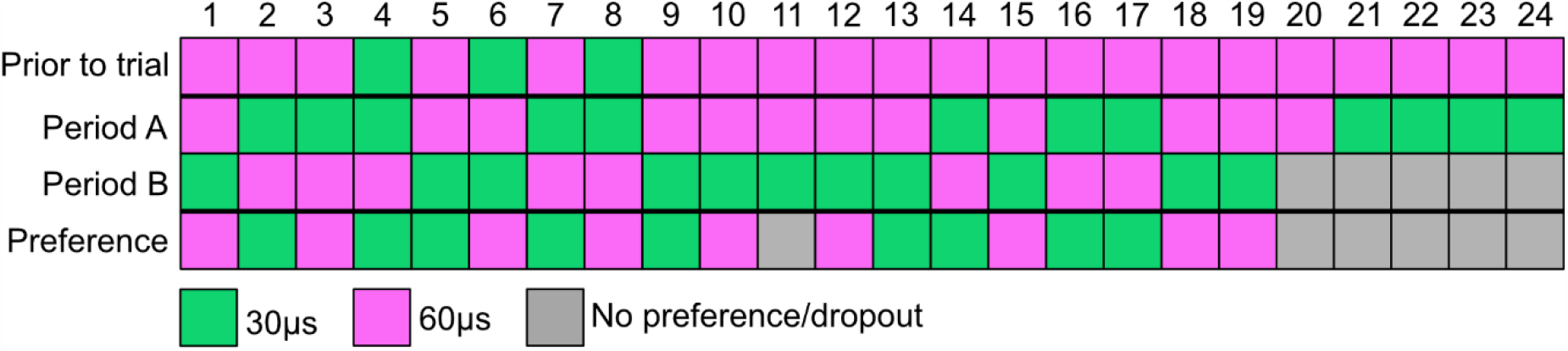
Individual Order of Pulse Widths. Before the trial a stimulation setting with 60µs was chosen in 21 of 24 patients. At the end of the trial 9 of 19 patients, who completed the trial,preferred a stimulation with 30µs or 60µs respectively.

### Clinical Outcomes

As shown in Table 1, there were no differences in time spent in the respective motor states as obtained by the motor diary between the PW60 and the PW30 setting. Additionally, there was no significant period-effect detected (data not shown). Furthermore, an analysis only including patients who completed both treatment conditions was conducted, demonstrating the same result (see. Supplementary Table 3). For secondary outcomes there were no differences between the two treatment conditions for UPDRS-I (PW60: 1.5 ±1.8, PW30: 1.3 ±1.7, p = 1.00), UPDRS-II (PW60: 8.7 ±3.4, PW30: 6.6 ±3.6, p = 0.1), UPDRS-III (PW60: 16.9 ±6.5, PW30: 16.1 ±6.6, p = 1.0), PDQ-39 SI (PW60: 64.1 ±47.3, PW30: 59.1 ±47.7, p = 1.0), and NMSQ (PW60: 7.6 ±3.6, PW30: 6.7 ±4.0, p = 1.00). Regarding side-effects there were no differences for VAS “ability to speak” (PW60: 4.8 ±2.9, PW30: 5.7 ±2.7, p = 1.00), intelligibility ratings (PW60: 8.0 ±3.6, PW30: 6.7 ±4.0, p = 1.00), VAS “ability to walk” (PW60: 5.4 ±2.8, PW30: 5.4 ±2.6, p = 1.00), and 10-m timed walk test (PW60: 8.6s ±1.6, PW30: 9.3 ±1.9, p = 0.18).

**Table 1.** Motor Diary Outcomes

| Motor States [h] | Period A<br>N = 24 |  | Period B<br>N = 19 |  | p | PW60<br>N = 20 |  | PW30<br>N = 23 |  | p |
| --- | --- | --- | --- | --- | --- | --- | --- | --- | --- | --- |
|  | Mean | SD | Mean | SD |  | Mean | SD | Mean | SD |  |
| „On“ | 11.95 | 4.34 | 10.65 | 4.92 | 1.0 | 11.7 | 4.07 | 11.09 | 5.08 | 1.0 |
| „Off“ | 1.57 | 3.19 | 2.14 | 3.71 | 1.0 | 1.92 | 3.6 | 1.73 | 3.29 | 1.0 |
| „Dyskinesia“ | 2.18 | 3.14 | 2.76 | 3.51 | 1.0 | 2.02 | 2.55 | 2.8 | 3.83 | 1.0 |
| „Sleep“ | 8.41 | 1.29 | 8.39 | 1.64 | 0.8 | 8.31 | 1.35 | 8.48 | 1.53 | 1.0 |

**Abbreviations:** SD = standard deviation, PW30 = trial condition with 30µs, PW60 = trial condition with 60µs.

### Energy Consumption

As shown in Figure 4, there were differences between the PW60 and PW30 setting, concerning CPP (PW60: 271.8nC ±118.86, PW30: 206.87nC ±103.67, p < 0.001) but not for TEED (PW60: 73.56µJ/s ±45.17, PW30: 92.22µJ/s ±66.46, p = 0.051) nor BCI (PW60: 38.88 ±20.98, PW30: 47.05 ±31.25, p = 0.123).

## Discussion

In this double-blind, crossover trial in patients with PD and bilateral STN-DBS, motor symptom control was not significantly different for short and conventional pulse widths. Additionally, there were no differences regarding any side-effect investigated, non-motor symptoms, quality-of-life, or energy consumption measures.

While most of the previous studies focused on an acute challenge – and often limited their analysis to differences in amplitude thresholds - the present study reports a comparison of motor symptom control after 4 weeks of continuous stimulation between the two pulse width conditions.^10–12^ In line with a previous study by Dayal et al.,^13^ our study provides evidence for a non-inferiority regarding motor symptom control and an acceptable safety profile of STN-DBS with a pulse width of 30µs. Unlike the present study, Dayal et al. only included patients with stimulation-induced dysarthria and thereby stimulation settings had already been adapted to this troublesome side-effect, which can lead to suboptimal motor symptom control. Additionally, in the present study, motor symptom control in each condition was evaluated over three days using a standardized motor diary, providing a home-based overall impression of motor symptom control under everyday conditions.^18^ This might especially be important in patients with advanced Parkinson’s disease, as motor fluctuations throughout the day could influence subjective motor outcomes and might not be captured by frequently used scales as, e.g., the UPDRS in a single visit.^2,18,19^

Subjectively, 47.5% of the patients (N = 9/19), tested in both conditions, preferred either the short (30µs) or the conventional (60µs) setting (see Figure 2). This individual preference has also been reported by Seger et al. regarding gait outcomes in short versus conventional pulse width settings.^16^ Although the group size did not allow for distinct subgroup analysis the results presented here point towards different patient profiles regarding preferences of pulse width settings. While patients preferring 30µs spent more time with troublesome dyskinesia in their non-preferred setting, patients preferring 60µs experienced more time with bradykinesia throughout the day (see Figure 3C and Supplementary Table 4). This concept is supported by a recent study by Dayal et al. demonstrating an alleviation of dyskinesia by stimulation with 30µs in an acute challenge.^15^

**Fig. 3.**
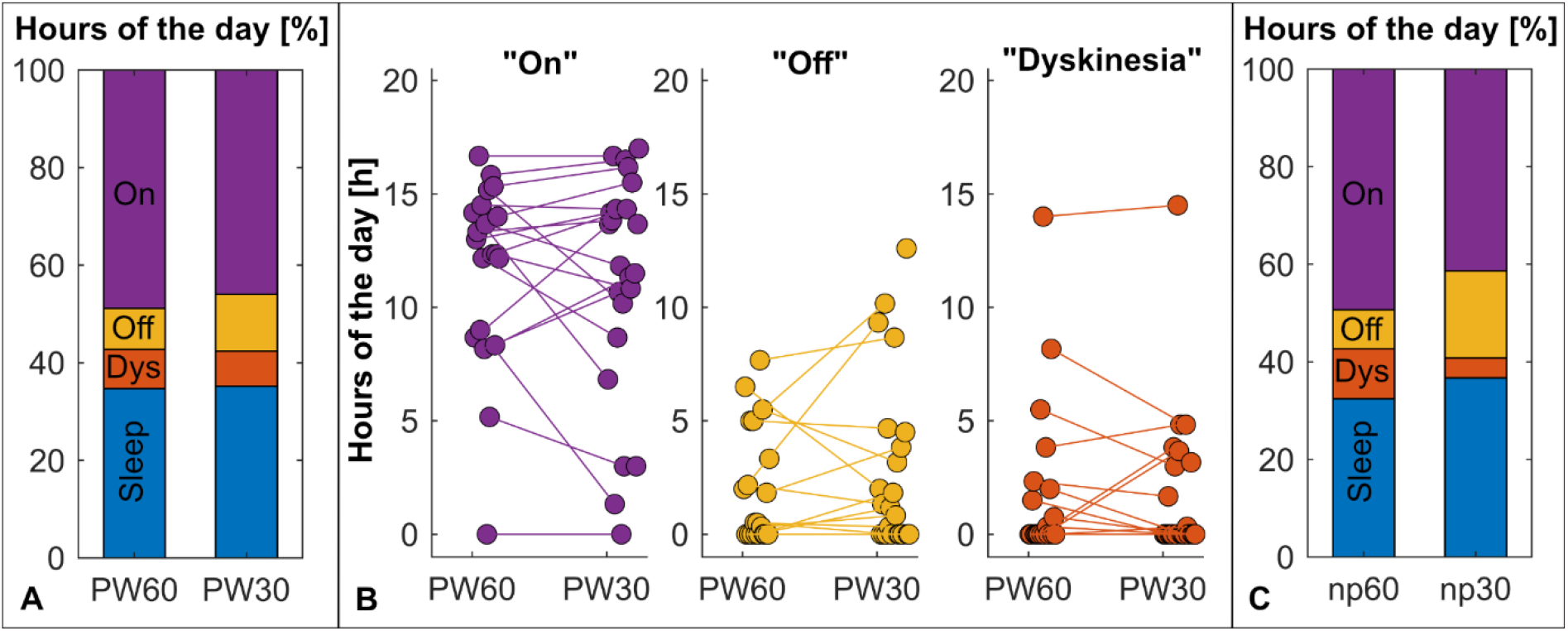
Motor Diary Outcome. (A) There were no differences in motor states as on the group level between treatment conditions. (B) On the subject level, individual differences between the treatment conditions were observed. (C) Patients preferring 30µs (N = 9) seemed to experience more dyskinesia in their non-preferred 60µs setting (np60), whereas patients preferring 60µs (N = 9) tend to have more off-time in their 30µs setting (np30). **Abbreviations:** np30 = 30µs condition of patients preferring 60µs, np60 = 60µs condition of patients preferring 30µs, PW30 = trial condition with 30µs, PW60 = trial condition with 60µs.

**Fig. 4.**
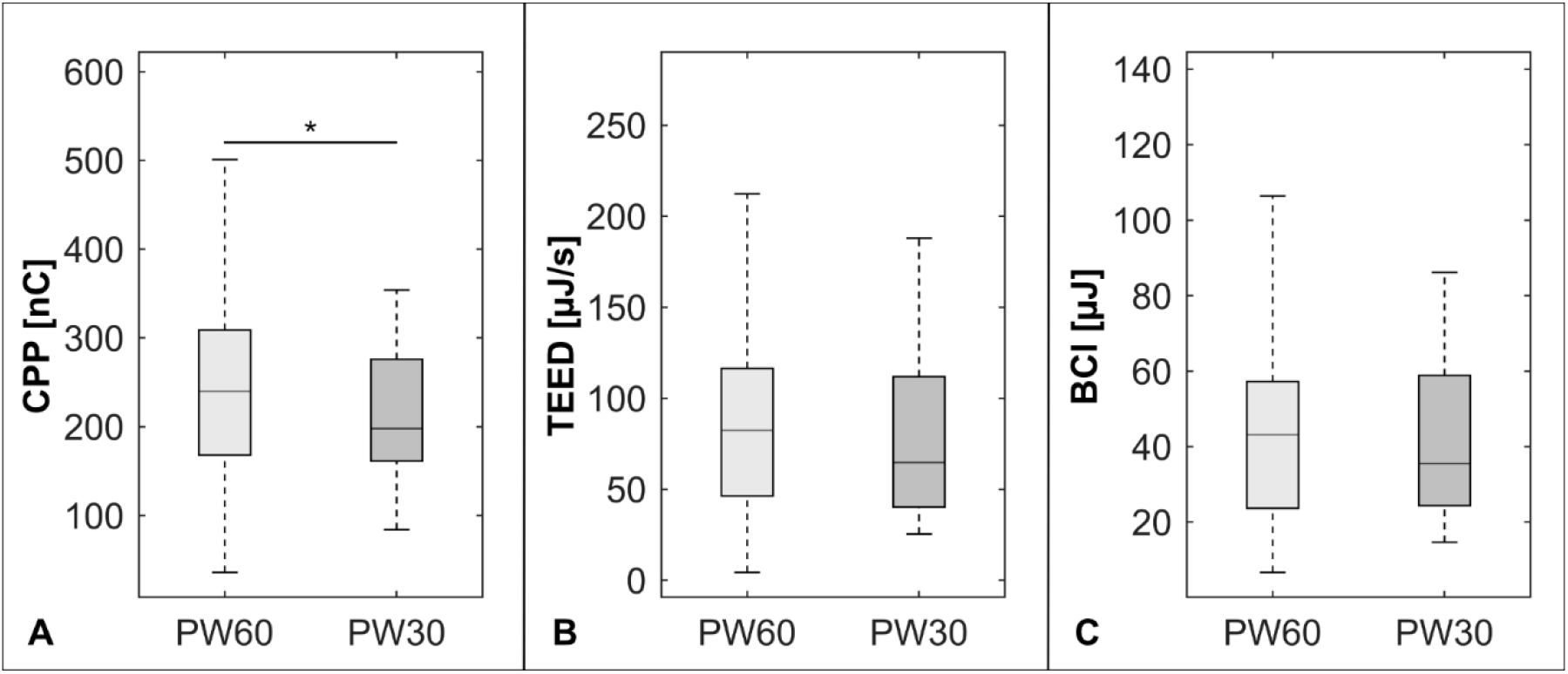
Energy Indices. (A) The 30µs condition resulted in a lower charge per pulse, whereas the total electrical energy delivered (B), and the battery drain was equal between the two conditions (C). (*p<0.001) **Abbreviations:** BCI = Battery Charge Index, CPP = charge per pulse, PW30 = trial condition with 30µs, PW60 = trial condition with 60µs, TEED = total electrical energy delivered.

An often-discussed benefit of short pulse width stimulation is its reduced charge per pulse. Previous studies hypothesized, that this might result in fewer side-effects and increased battery life.^10–12^ The potential benefit of preventing stimulation-induced side effects has mainly been derived from an increase of the therapeutic window in terms of a widening of the amplitude range between side-effect and efficacy threshold.^11–14,22^ Beneficial effects of short pulse width stimulation on side-effects have only been reported in DBS for essential tremor patients,^23^ but not in patients with PD beyond the setting of an acute challenge.^13,14^ Although Reich et al. concluded that a wider therapeutic window might result from selectivity of short pulse widths for smaller, beneficial axons over larger, side-effect associated fibers,^10^ this hypothesis has been falsified in prior and current studies.^22,24^ Instead, stimulation with shorter pulse widths may lead to a preferential activation of larger axons with shorter chronaxies, as demonstrated by Anderson et al.^22^ Regarding battery life, it has to be clarified that it critically depends on energy consumption and not only charge. Therefore we calculated the TEED and the BCI, an energy index also used to indicate energy consumption of stimulation settings on the programming device per clinical routine.^21^ Both energy consumption measures were equal for the two conditions. This demonstrates that stimulating with 30µs has no beneficial effect on battery life and is in line with a previous study reporting equivalency in TEED when comparing both settings at the efficacy threshold in an acute challenge.^14^ It also highlights the important fact, that when only taking one of those parameters into account, e.g. amplitude, charge, TEED or BCI, different results regarding its energy consumption can be obtained by definition when comparing 30µs to 60µs stimulation.

The main limitation of the present trial is that the calculated sample size of N = 27 to show non-inferiority of short pulse width stimulation with a power of 80% has not been reached due to an unexpected high number of dropouts during the trial. Nevertheless, the sample size is one of the biggest in studies investigating the effect of short pulse widths in STN-DBS. These dropouts reflect the difficulty of conducting a clinical trial during the COVID-19 pandemic, which is why visits in between the follow-up visits were kept to a minimum and patients were mainly supervised via telephone. Additionally, the possibility to change stimulation settings via the patient programmer was another source of dropouts, as two patients switched back to the previous condition’s setting instead of titrating the amplitude. However, in line with our clinical routine, we discharged patients with a well-established “backup” stimulation setting for patient safety reasons. Nevertheless, the study size seems sufficient as not even a trend for a difference in motor symptom control between the study conditions was observed despite inclusion of more than 80% of the calculated sample size (all p = 1.0, see table 1). Of note, due to the dropouts, the planned 1:1 randomization was not achieved, but no period-effect was observed. Secondly, the choice of the stimulation settings was based on a single non-standardized clinical programming session instead of a standardized monopolar review as it has been part of a similar trial by Dayal et al.^13^ As mentioned before, Dayal et al. included patients with stimulation-induced dysarthria in contrast to the present study. Therefore, every patient needed an elaborated adjustment of the stimulation setting before study inclusion, which was not the case in the present cohort. Another difference to prior studies is that mainly patients themselves titrated the amplitude. To prevent potentially suboptimal stimulation settings,^25^ patients were well trained at baseline and reinstructed per protocol telephone calls if motor symptom control was insufficient or side-effects were reported. Overall real-life outcomes may even be better reflected by this practice, as it is unlikely that patients present to an outpatient clinic for stimulation adjustment regularly within such a short time frame.

To conclude, the present study provides evidence that there is no difference concerning motor symptom control, the occurrence of side-effects or energy consumption between short and conventional pulse width stimulation in STN-DBS for patients with PD on the group level when tested under real-life conditions. However, future studies are warranted with larger patient samples to investigate whether patients with distinct symptom burden profiles, especially dyskinesia, might benefit from shorter pulse width stimulation.

## Data Availability

Data is available upon reasonable request to the authors.

## Acknowledgment

We thank our patients for contributing to this study.

## Authors’ Roles

1. Research project: A. Conception, B. Organization, C. Execution;
2. Statistical Analysis: A. Design, B. Execution, C. Review and Critique;
3. Manuscript: A. Writing of the first draft, B. Review and Critique.

JNPS: 1A, 1B, 1C, 2A, 2B, 3A; LMS: 1B, 1C, 2C, 3B; HJ: 1C, 2C, 3B; PR: 1A, 1C, 2C, 3B, JKS: 1C, 2C, 3B, HSD: 2C, 3B; JCB: 2C, 3B; GRF: 2C, 3B; VVV: 2C, 3B; TAD: 1A, 2C, 3B; MTB: 1A, 2C, 3B.

## Abbreviations

BCI: Battery Charge Index,
CPP: Charge per pulse,
NMSQ: Non-motor symptoms questionnaire,
PD: Parkinson’s disease,
PDQ-39 SI: Parkinson’s disease questionnaire 39-Item Quality of Life Questionnaire Summary Index,
PW30: trial condition with 30µs,
PW60: trial condition with 60µs,
STN-DBS: subthalamic nucleus deep brain stimulation,
TEED: total electrical energy delivered,
UPDRS: Unified Parkinson’s disease rating scale,
VAS: Visual analog scale

## Financial Disclosures of all authors (for the preceding 12 months)

Jan Niklas Petry-Schmelzer has nothing to report.

Lisa M Schwarz has nothing to report.

Hanna Jergas has nothing to report.

Paul Reker has nothing to report

Julia K. Steffen has nothing to report.

Haidar S. Dafsari was funded by the EU Joint Programme – Neurodegenerative Disease Research (JPND), the Prof. Klaus Thiemann Foundation in the German Society of Neurology, the Felgenhauer Foundation, the KoelnFortune program of the Medical Faculty of the University of Cologne, and has received honoraria by Boston Scientific, Medtronic and Stadapharm.

Juan Carlos Baldermann has nothing to report.

Gereon R. Fink serves as an editorial board member of Cortex, Neurological Research and Practice, NeuroImage: Clinical, Zeitschrift für Neuropsychologie, and DGNeurologie; receives royalties from the publication of the books Funktionelle MRT in Psychiatrie und Neurologie, Neurologische Differentialdiagnose, and SOP Neurologie; receives royalties from the publication of the neuropsychological test batteries KAS, Köpps, and NP-Kiss; received honoraria for speaking engagements from DGN and Forum für medizinische Fortbildung FomF GmbH.

Veerle Visser-Vandewalle received speaker’s honoraria by Boston Scientific, Medtronic and LivaNova.

Till A. Dembek has nothing to report.

Michael T. Barbe received speaker’s honoraria from Medtronic, Boston Scientific, Abbott (formerly St. Jude), GE Medical, UCB, Apothekerverband Köln e.V. and Bial as well as research funding from the Felgenhauer-Stiftung, Forschungspool Klinische Studien (University of Cologne), Horizon 2020 (Gondola), Medtronic (ODIS), and Boston Scientific and advisory honoraria for the IQWIG.

## Supplementary

**Supplementary Table 1.** Adverse Events. Adverse events as reported at follow-up visits or at dropout.

| Event | Number of AE<br>in PW60 | Number of AE<br>in PW30 | Total number of subjects<br>in both conditions |
| --- | --- | --- | --- |
| *Adverse Events associated with stimulation* |  |  |  |
| Dyskinesia | 8 | 8 | 9 |
| Speech impairment | 2 | 1 | 3 |
| Worsening of symptom control | 1 | 3 | 4 |
| **Total** | **11** | **12** | **13/30** |
| *Adverse Events probably associated with stimulation* |  |  |  |
| Numbness in a limb | 1 | 0 | 1 |
| Spasms in fingers or toes | 2 | 1 | 2 |
| Impairment of balance | 1 | 0 | 1 |
| **Total** | **4** | **1** | **4/30** |
| *Adverse Events not associated with stimulation* |  |  |  |
| Neckpain | 1 | 1 | 1 |
| Pain in a limb | 2 | 1 | 3 |
| Dorsal pain | 0 | 2 | 2 |
| Mild depressive symptoms | 5 | 3 | 5 |
| Disrupted sleep | 0 | 1 | 1 |
| **Total** | **8** | **8** | **11/30** |
| *Serious Adverse Event not associated with stimulation* |  |  |  |
| DBS system infection | 1 | 0 | **1/30** |

**Abbreviations:** AE = adverse event, DBS = deep brain stimulation, PW30 = trial condition with 30µs, PW60 = trial condition with 60µs.

**Supplementary Table 2.** Stimulation Settings at Follow-up Visits.

| Subject | Stimulation Setting | Amplitude<br>PW60 [mA] | Amplitude<br>PW30 [mA] |
| --- | --- | --- | --- |
| 1 | R: C+, 3- (100%), 119 Hz<br>L: C+, 9- (25%), 10- (25%), 11- (25%), 119 Hz | R: 1.8<br>L: 8.1 | R: 3.7<br>L: 8.4 |
| 2 | R: C+, 1- (10%), 2- (10%), 3- (10%), 4- (24%), 5- (23%), 6- (23%), 130 Hz<br>L: C+, 9- (10%), 10- (10%), 11- (10%), 12- (24%), 13- (23%), 14- (23%), 130 Hz | R: 2.7<br>L: 2.3 | R: 4.0<br>L: 4.0 |
| 3 | R: C+, 0- (60%), 1- (20%), 2- (20%), 130 Hz<br>L: C+, 8- (60%), 9- (18%), 10- (22%), 130 Hz | R: 1.8<br>L: 2.1 | R: 2.9<br>L: 3.1 |
| 4 | R: C, 1+ (50%), 2- (100%), 3+ (50%), 130 Hz<br>L: C+, 9- (30%), 11- (30%), 12- (40%), 174 Hz | R: 2.0<br>L: 1.7 | R: 3.0<br>L: 3.0 |
| 5 | R: C+, 1- (34%), 2- (33%), 3- (33%), 116 Hz<br>L: C+, 11- (50%), 14- (50%), 116 Hz | R: 2.2<br>L: 3.1 | R: 4.3<br>L: 5.0 |
| 6 | R: C+, 1- (34%), 2- (33%), 3- (33%), 130 Hz<br>L: C+, 9- (34%), 10- (33%), 11- (33%), 130 Hz | R: 3.0<br>L: 3.4 | R: 4.3<br>L: 4.9 |
| 7 | R: C+, 1- (34%), 2- (33%), 3- (33%), 130 Hz<br>L: C+, 12- (34%), 13- (33%), 14- (33%), 130 Hz | R: 2.5<br>L: 1.5 | R: 4.0<br>L: 2.8 |
| 8 | R: C+, 7- (100%), 130 Hz<br>L: C+, 15- (100%), 130 Hz | R: 0.2<br>L: 1.2 | R: 0.2<br>L: 1.0 |
| 9 | R: C+, 4- (34%), 5- (33%), 6- (33%), 130 Hz<br>L: C+, 12- (34%), 13- (33%), 14- (33%), 130 Hz | R: 0.7<br>L: 0.7 | R: 1.1<br>L: 0.7 |
| 10 | R: C+, 4- (34%), 5- (33%), 6- (33%), 119 Hz<br>L: C+, 12- (50%), 13- (50%), 119 Hz | R: 2.9<br>L: 2.9 | R: 5.8<br>L: 6.3 |
| 11 | R: C+, 4- (100%), 130 Hz<br>L: C+, 9- (100%), 130 Hz | R: 0.7<br>L: 1.9 | R: 1.4<br>L: 3.3 |
| 12 | R: C+, 4- (33%), 5- (34%), 6- (33%), 130 Hz<br>L: C+, 12- (10%), 13- (10%), 14- (10%), 15- (70%), 130 Hz | R: 2.1<br>L: 2.6 | R: 4.6<br>L: 4.5 |
| 13 | R: C+, 1- (16%), 2- (16%), 4- (33%), 5- (35%), 185 Hz<br>L: C+, 10- (13%), 11- (15%), 13- (36%), 14- (36%), 185 Hz | R: 3.2<br>L: 2.7 | R: 4.9<br>L: 3.8 |
| 14 | R: C+, 4- (31%), 5- (42%), 6- (27%), 130 Hz<br>L: C+, 12- (42%), 13- (42%), 14- (16%), 130 Hz | R: 2.3<br>L: 2.3 | R: 3.3<br>L: 3.3 |
| 15 | R: C+, 4- (34%), 5- (33%), 6- (33%), 130 Hz<br>L: C+, 12- (34%), 13- (33%), 14- (33%), 130 Hz | R: 2.2<br>L: 2.4 | R: 2.8<br>L: 3.3 |
| 16 | R: C+, 3- (100%), 204 Hz<br>L: C+, 12- (34%), 13- (33%), 14- (33%), 204 Hz | R: 2.7<br>L: 0.8 | R: 4.0<br>L: 1.0 |
| 17 | R: C+, 4- (33%), 5- (34%), 6- (33%), 198 Hz<br>L: C+, 12- (34%), 13- (33%), 14- (33%), 198 Hz | R: 1.5<br>L: 1.3 | R: 2.6<br>L: 1.9 |
| 18 | R: C+, 1- (34%), 2- (33%), 3- (33%), 130 Hz<br>L: C+, 9- (50%), 10- (50%), 130 Hz | R: 1.2<br>L: 1.9 | R: 2.4<br>L: 2.9 |
| 19 | R: C+, 1- (18%), 2- (16%), 3- (16%), 4- (18%), 5- (16%), 6- (16%), 130 Hz<br>L: C+, 12- (34%), 13- (33%), 14- (33%), 130 Hz | R: 2.6<br>L: 2.6 | R: 3.4<br>L: 3.1 |
| 20 | R: C+, 2- (34%), 3- (33%), 4- (33%), 130Hz<br>L: C+, 10- (34%), 11- (33%), 12- (33%), 130Hz | R: 2.4<br>L: 2.9 | R: n.a.<br>L: n.a. |
| 21 | R: C+, 4- (100%), 130Hz<br>L: C+, 9- (100%), 130Hz | R: n.a.<br>L: n.a. | R: 1.1<br>L: 1.7 |
| 22 | R: C+, 5- (34%), 6- (33%), 7- (33%), 179Hz | R: n.a. | R: 4.6 |
|  | L: C+, 13- (50%), 15- (50%), 179Hz | L: n.a. | L: 6.4 |
| 23 | R: C+, 5- (34%), 6- (33%), 7- (33%), 130Hz | R: n.a. | R: 3.1 |
|  | L: C+, 13- (34%), 14- (33%), 15- (33%), 130Hz | L: n.a. | L: 4.0 |
| 24 | R: C+, 5- (34%), 6- (33%), 7- (33%), 174Hz | R: n.a. | R: 0.6 |
|  | L: C+, 10- (18%), 11- (16%), 12- (16%), 13- (18%), 14- (16%), 15- (16%), 174Hz | L: n.a. | L: 3.5 |

Abbreviations: C = Case, L = left hemisphere, n.a. = not available due to dropout; PW30 = trial condition with 30 μs, PW60 = trial condition with 60 μs, R = right hemisphere.

**Supplementary Table 3.**
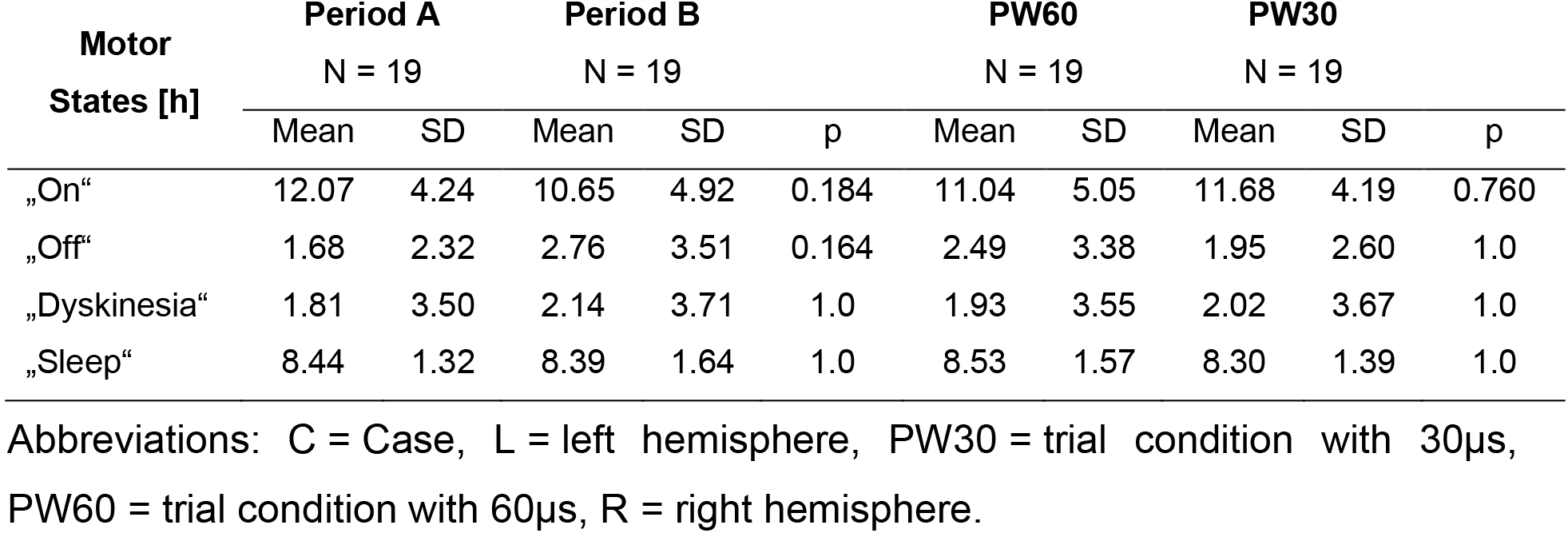
Motor Diary Outcomes for paired data only.

**Supplementary Table 4.** Subgroup analysis: Comparison of non-preferred settings.

|  | npPW60 (N = 9) |  | npPW30 (N = 9) |  | p |
| --- | --- | --- | --- | --- | --- |
|  | Mean | SD | Mean | SD |  |
| *Motor states [h]* |  |  |  |  |  |
| „On“ | 11.78 | 4.20 | 9.94 | 4.47 | 1.0 |
| „Off“ | 1.91 | 3.03 | 4.27 | 3.70 | 0.436 |
| „Dyskinesia“ | 2.46 | 2.89 | 0.98 | 1.71 | 0.864 |
| „Sleep“ | 7.74 | 1.20 | 8.81 | 2.04 | 0.988 |
| *Secondary outcomes* |  |  |  |  |  |
| VAS „ability to speak“ | 4.13 | 3.09 | 5.69 | 2.14 | 0.928 |
| Intelligibility | 8.25 | 1.39 | 7.76 | 1.11 | 1.00 |
| VAS „ability to walk“ | 5.03 | 2.82 | 5.08 | 2.42 | 1.00 |
| 10-m timed walk [s] | 8.13 | 1.45 | 9.78 | 1.22 | 0.076 |
| UPDRS-I | 1.22 | 1.20 | 1.89 | 2.37 | 1.00 |
| UPDRS-II | 9.00 | 3.46 | 7.67 | 3.12 | 1.00 |
| UPDRS-III | 19.56 | 6.91 | 16.89 | 6.33 | 1.00 |
| PDQ39-SI | 22.89 | 15.62 | 27.22 | 24.97 | 1.00 |
| NMSQ | 7.22 | 3.15 | 7.78 | 5.17 | 1.00 |
| *Energy indices* |  |  |  |  |  |
| Battery charge index [μJ] | 43.56 | 21.92 | 44.66 | 33.62 | 1.00 |
| Charge per pulse [nC] | 241.33 | 82.69 | 240.67 | 133.55 | 1.00 |
| Total electrical energy delivered [μJ/s] | 83.60 | 48.83 | 90.32 | 78.72 | 1.00 |

**Abbreviations:** npPW60 = 60μs condition of patients preferring 30μs setting, npPW30 = 30μs condition of patients preferring 60μs setting, SD = standard deviation

## Notes

**Financial Disclosure/Conflict of Interest concerning the research related to the manuscript:** None

**Funding sources for the study:** JNPS and TAD were supported by the Cologne Clinician Scientist Program (CCSP)/Faculty of Medicine/University of Cologne. Funded by the German Research Foundation (DFG, FI 773/15-1). JCB and VVV were funded by the Deutsche Forschungsgemeinschaft, German Research Foundation (Project-ID: 431549029 – SFB 1451).

### Competing Interest Statement

The authors have declared no competing interest.

### Clinical Trial

German Clinical Trials Register No. DRKS00017528

### Funding Statement

No external funding.

### Author Declarations

The ethics committee of the University of Cologne approved the trial (vote: 19-1233) and it was conducted under the declaration of Helsinki.

